# Sexual orientation disparities in depression and substance use among adults: Results from the Brazilian National Health Survey, 2019

**DOI:** 10.1101/2022.06.18.22275833

**Authors:** Nayara L. Gomes, Claudia S. Lopes

**Affiliations:** Social Medicine Institute, Department of Epidemiology, Rio de Janeiro State University, Rio de Janeiro, Brazil; Brazilian Institute of Geography and Statistics, Department of Study, Methods and Control, Coordination of Household Sample Surveys, Rio de Janeiro, Brazil

**Keywords:** Sexual orientation, Health disparities, Depression, Substance use/Abuse, Adults, Brazil

## Abstract

**Purpose:** To compare the prevalence of depression and substance use/abuse according to sexual orientation and sex among Brazilian adults.

**Methods:** Data were obtained from a national health survey (PNS 2019). This study included participants aged 18 and over (N=85,859). Prevalence ratios were estimated using Poisson models stratified by sex, for the association between sexual orientation, depression, daily tobacco use, alcohol abuse and both substance use/abuse.

**Results:** After controlling for covariates, gay men showed a higher prevalence of depression (APR=1.92, 95% CI 1.32; 2.80), daily tobacco use (APR=1.93, 95% CI 1.45; 2.56) and multiple substances use/abuse (APR=1.73, CI95% 1.14;2.62) than heterosexuals. Bisexual men showed higher prevalence of depression (APR=2.91, 95% CI 1.54; 5.53). Estimated prevalence ratios were even higher among women. Lesbians showed higher prevalence of binge drinking (APR=2.52, 95% CI 1.94; 3.27), heavy drinking (APR=3.36, 95% CI 2.39; 4.72), daily tobacco use (APR=2.66, 95% CI 1.74; 4.07) and multiple substances use/abuse (APR=4.35, 95% CI 2.35;8.07) than among heterosexuals. Among bisexual women, results were significant for all analyzed outcomes: depression (APR=2.32, 95% CI 1.72; 3.13), binge drinking (APR=2.52, 95% CI 1.94; 3.27), heavy drinking (APR=2.53, 95% CI 1.68; 3.81), daily tobacco use (APR=1.82, 95% CI 1.12;2.96) and multiple substances use/abuse (APR=3.23, 95% CI 1.72;6.05).

**Conclusions:** Our results indicate that sexual minorities are more vulnerable to mental illness and substance use/abuse. This finding draws attention to the need for specific public policies aimed at this population and for a greater recognition and better management of these disorders by health professionals.

## Introduction

The non-heteronormative part of the population has been identified as more vulnerable to certain negative health outcomes. Several authors have been pointing to evidence of health disparities among Lesbians, Gays and Bisexuals (LGB) compared to their heterosexual counterparts. Among them are those related to mental health or the use of substances such as alcohol, cigarettes and illicit drugs ^1-4^. Some studies have documented more pronounced disparities within specific subgroups of the population as among women ^5-9^ or bisexual ^1, 5,10^.

The theory known as “minority stress model” is recurrently used to explain such sexual orientation-related disparities, suggesting that certain conditions and structures of the social environment end up acting as sources of stress, leading to harmful mental and physical effects and having a strong impact on the lives of people who belong to stigmatized minority groups ^11,12^. Discrimination suffered by sexual minority individuals, for example, would be one of the risk factors for negative health outcomes in this subgroup of the population ^13-17^.

There is a growing demand for the inclusion of sexual orientation in population surveys to enable the monitoring of disparities related to this aspect ^18^. Although there is already an increasing literature on health issues of the LGB population, there is a concentration of studies conducted in the United States and in European countries ^9,19-23^.

In Brazil, the country where the higher number of LGBT people are killed in the world ^24^, and where religious intolerance has been criticized for contributing to stigma and to a pathological view of this population group ^23^, there is still a lack of data on the development of public policies, despite the existence of a national comprehensive health policy for Lesbians, Gays, Bisexuals, Transvestites and Transsexuals, established in 2011^25^.

According to a recent systematic review, although Brazil is the Latin American country where we find almost all the scientific production on the health of the LGBT population in the region (in addition to the topic of HIV/AIDS), studies are still scarce, having been increasing in number only since 2014 and being mostly qualitative or focused on professional practice, mental health and assessment of programs ^26^.

Population-based surveys, using probabilistic samples, which simultaneously address the sexual orientation and health of the population are scarce in countries such as Brazil. Existing studies dealing with this topic often involve convenience samples, focuses on specific regions of the country, or only include LGB people ^21,27,28^. On the topic of mental health and substance use, the only national-level study identified examined only substance use without stratification by sex ^23^.

The data collected in the 2019 National Health Survey, the most comprehensive population health survey in Brazil, included, for the first time, the investigation of sexual orientation among the adult population in the national territory, enabling the analysis of differential health outcomes according to sexual orientation of individuals.

The aim of our study was to compare the prevalence of use/abuse of substances, such as alcohol and tobacco, as well as of depression, across the different sexual orientations as self-reported by adults in Brazil, according to sex.

## Methods

### Participants and procedures

The data used in this study refer to the 2nd edition of the National Health Survey (PNS-2019) conducted by Brazilian Institute of Geography and Statistics (IBGE) in partnership with Ministry of Health. It is a nationwide household survey with a three-stage cluster sampling design ^29^. Sexual orientation question was included for the first time in this edition, applied only to selected residents aged 18 and over ^30^.

Our study included 85,859 individuals who self-identified as heterosexual (84,359), homosexual or bisexual (n=1,500).

People who reported other sexual orientations (n=54), or who did not know their sexual orientation (n=849), and those who refused to answer this question (n=1,769) were not considered.

The PNS was approved by the National Research Ethics Committee (Process: 3,529,376, of August 23, 2019). The consent of the participants was obtained in two stages, the first at the beginning of the household interview and the second at the interview of the selected resident ^29^.

### Measures

#### Sexual orientation

Self-identified sexual orientation was investigated, through the question “What is your sexual orientation?” and alternatives: 1. Heterosexual; 2. Bisexual; 3. Homosexual; 4. Other; 5. Does not know; 6. Refused to answer.

In this study we compared heterosexuals with bisexuals and homosexuals.

#### Abusive alcohol use

Abusive alcohol use was assessed using two different use patterns.

Those who answered “Yes” to the question “In the last thirty days, have you consumed five or more doses of alcoholic beverages on a single occasion?” were classified as reporting binge drinking.

To assess heavy drinking, the following questions were used: “How many days a week do you usually consume an alcoholic beverage?” and “In general, on the day you drink, how many doses of alcohol do you consume?” In our study, heavy drinking was considered as the weekly consumption of 8 drinks or more (for women) and 15 drinks or more (for men), according to CDC criteria ^31^.

#### Daily tobacco use

The daily use of cigarettes or any other tobacco product, smoked or not – such as straw or hand-rolled cigarettes, clove or Bali cigarettes, pipes, cigarillos, hookah or water pipes, snuff, electronic cigarettes – was considered for those who answered “Yes, daily” to the question “Do you currently smoke any tobacco products?”

#### Use/abuse of multiple substances

In our study, the use/abuse of multiple substances refers to those who reported daily use of tobacco products while also reporting excessive alcohol use (binge drinking or heavy drinking).

#### Depression

Depression was investigated in the survey using the Patient Health Questionnaire-9 (PHQ-9), which assesses the frequency of depressive symptoms in the last two weeks prior to data collection.

In our study, the presence of major depressive disorders was defined by a PHQ-9 score equal to or higher than 10, which is considered the best cut-off point for this detection ^32^. In the specific case of the Brazilian adult population, a study found that ≥ 9 and ≥ 10 were the cutoff points that showed the highest sensitivity and specificity for the diagnosis of depression ^33^.

#### Covariates

The variables considered in the analyses, based on the literature, were: i) age groups: 18 to 24, 25 to 39, 40 to 59 and 60 years or older; ii) self-declared color or race: white, black, brown and others; iii) marital status: married or cohabiting with a partner and non-cohabiting single; iv) level of education: no schooling or some elementary school, elementary school or some high school, high school and some college or more; v) situation of the household, whether urban or rural;

### Statistical analysis

The characteristics of the study sample were summarized using totals and percentages. The chi-square test was used to assess the statistical significance of the differences between the different sexual orientations’ characteristics analyzed.

The prevalence and respective 95% confidence intervals of the outcomes of interest were also calculated, according to sex and sexual orientation.

Poisson regression models with robust variance ^34^ were used to estimate crude and adjusted prevalence ratios and their respective 95% confidence intervals, to assess the association between sexual orientation and the use/abuse of substances and depression. All outcomes assessed in this study were analyzed with separate regression models stratified by sex.

The analyzes were performed using the survey commands (svy) of the Stata software version 14, used for data collected with surveys with a complex sample design. Therefore, all results presented considered the survey design and sample weights.

## Results

In 2019, based on National Health Survey data, there were 2.8% (95% CI: 1.7;2.0) of population aged 18 years and over who self-identified as homosexual or bisexual in Brazil, of which 0.9% self-identified as bisexuals, 0.7% as lesbians and 0.8% as gays.

Among men, the percentage of homosexuals (1.4%) was higher than that observed among women (0.9%), while the percentage of bisexuals was lower (0.5% and 0.8% for men and women, respectively).

The socioeconomic characteristics of the survey participants, according to sex and self-identified sexual orientation, are shown in table 1. We can observe that the proportion of young participants are higher among homosexuals and bisexuals compared to heterosexual. Among bisexual women, for example, 43.9% are aged 18 to 24 against 8.5% among heterosexual women.

**Table 1:** Socioeconomic characteristics of survey participants by sex and sexual orientation (N=85,859)

| Variables | Men |  |  |  | Women |  |  |  |
| --- | --- | --- | --- | --- | --- | --- | --- | --- |
|  | Heterosexual<br>(n=39,751) | Homosexual<br>(n=586) | Bisexual<br>(n=194) | p<sup>a</sup> | Heterosexual<br>(n=44,608) | Homosexual<br>(n=394) | Bisexual<br>(n=326) | p<sup>a</sup> |
| Age groups |  |  |  |  |  |  |  |  |
| 18 - 24 | 8.7 | 20.5 | 29.9 | <0.001 | 8.5 | 21.1 | 43.9 | <0.001 |
| 25 - 39 | 29.1 | 43.5 | 37.1 |  | 27.8 | 47.5 | 38.3 |  |
| 40 - 59 | 37.2 | 30.4 | 25.3 |  | 36.5 | 27.4 | 14.7 |  |
| 60 and over | 25.1 | 5.6 | 7.7 |  | 27.2 | 4.1 | 3.1 |  |
| Color or race |  |  |  |  |  |  |  |  |
| White | 36.4 | 40.1 | 28.9 | 0.537 | 37.0 | 41.6 | 38.7 | 0.494 |
| Black | 11.8 | 12.1 | 11.9 |  | 11.1 | 10.2 | 13.2 |  |
| Mixed race | 50.3 | 46.4 | 55.7 |  | 50.5 | 47.0 | 44.8 |  |
| Others | 1.5 | 1.4 | 3.6 |  | 1.5 | 1.3 | 3.4 |  |
| Marital status |  |  |  |  |  |  |  |  |
| Single noncohabiting | 33.7 | 76.3 | 84.0 | <0.001 | 49.1 | 57.9 | 77.6 | <0.001 |
| Married or cohabiting | 66.3 | 23.7 | 16.0 |  | 50.9 | 42.1 | 22.4 |  |

| Education level |  |  |  |  |  |  |  |  |
| --- | --- | --- | --- | --- | --- | --- | --- | --- |
| No schooling or<br>some elementar school | 43.4 | 12.0 | 11.9 | <0.001 | 38.2 | 11.4 | 9.2 | <0.001 |
| Elementary school<br>or some high school | 14.1 | 10.2 | 11.3 |  | 12.8 | 14.7 | 13.5 |  |
| High school | 25.1 | 28.2 | 33.5 |  | 27.2 | 34.5 | 31.0 |  |
| Some college or<br>more | 17.4 | 49.7 | 43.3 |  | 21.7 | 39.3 | 46.3 |  |
| Household status |  |  |  |  |  |  |  |  |
| Urban | 72.4 | 91.3 | 88.1 | 0.001 | 81.0 | 94.2 | 94.2 | 0.003 |
| Rural | 27.6 | 8.7 | 11.9 |  | 19.0 | 5.8 | 5.8 |  |

<sup>a</sup> P-value resulting of Pearson chi-square test for the association between sexual orientation and socioeconomic characteristics considering weighted proportions and survey design.

With regard to marital status, single people, or people who are not cohabiting with a partner, predominate among homosexuals or bisexuals in the sample, in contrast to observed among heterosexuals; the percentages of people with some college or more and from urban areas higher among homosexuals or bisexuals than among heterosexuals.

Color or race was the only variable that did not show a significant difference in distribution between sexual orientations for both sexes in the study sample. For this reason, was not considered a confounder variable and was not included as a control variable in the regression models.

For both sexes and the different sexual orientations, the prevalence of depression, abusive alcohol use, daily tobacco use and use/abuse of both substance were also calculated (Figure 1).

**Figure 1.**
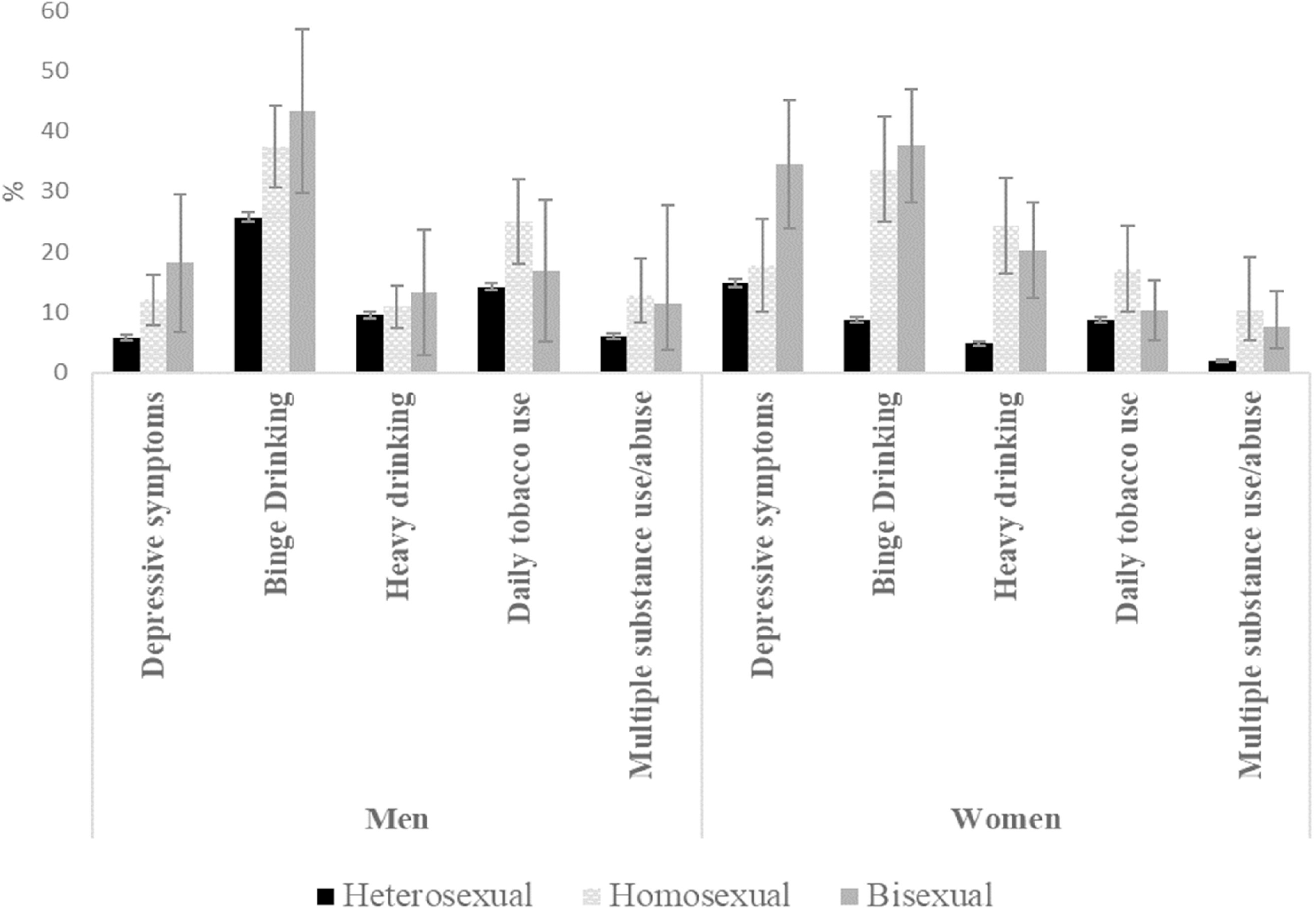
Depression and substance use/abuse prevalence and 95% confidence intervals

For all outcomes analyzed for both sexes, except for heavy drinking among men, we observe, based on the confidence intervals obtained, that there are significant differences in the estimated prevalence across the different sexual orientations considered.

Among men, we verified that the prevalence of depression between homosexual and bisexual was, respectively, more than double (12.0%) and triple (18.1%) of the prevalence observed among heterosexuals (5.7%). With regard to substance use, the observed daily use of tobacco and the use/abuse of multiple substances was higher among homosexual (25% and 12,7%, respectively) compared with heterosexual (14.2% and 5.9%). Binge drinking was the most prevalent outcome, being significatively higher between homosexual (37.4%) and bisexual (43.2%) compared with heterosexual (25.7%).

With respect to results for women, we found significant differences between lesbians or bisexuals compared with heterosexuals, for all outcomes considered. Especially noteworthy is the high percentage of bisexual women with depression (34.5%) and who reported binge drinking (37.5%).

Crude prevalence ratios observed for the total population aged 18 and over were significant for all outcomes under analysis when comparing homosexuals and bisexuals with heterosexuals, except for daily tobacco use among bisexuals (Table 2). All these results remain significant in adjusted models, confirming the higher prevalence of substance use/abuse and depression among sexual minorities, even after controlling for other important variables. Among the outcomes analyzed, depression among bisexuals, in comparison with heterosexuals, was that which showed the highest prevalence ratio (APR=2.47, 95% CI: 1.88;3.25).

**Table 2.** Prevalence ratios for the association of sexual orientation with depression and substance use/abuse

|  | Total |  |  |  | Women |  |  |  | Men |  |  |  |
| --- | --- | --- | --- | --- | --- | --- | --- | --- | --- | --- | --- | --- |
|  | CPR | 95% CI | APR | 95% CI | CPR | 95% CI | APR | 95% CI | CPR | 95% CI | APR | 95% CI |
| **Depression** |  |  |  |  |  |  |  |  |  |  |  |  |
| Bisexual | **2.73** | **[2.04,3.65]** | **2.47** | **[1.88,3.25]** | **2.32** | **[1.70,3.18]** | **2.32** | **[1.72,3.13]** | **3.15** | **[1.66,5.95]** | **2.91** | **[1.54,5.53]** |
| Homosexual | **1.37** | **[1.02,1.84]** | **1.52** | **[1.14,2.02]** | 1.20 | [0.77,1.86] | 1.22 | [0.79,1.89] | **2.09** | **[1.46,2.99]** | **1.92** | **[1.32,2.80]** |
| **Binge Drinking** |  |  |  |  |  |  |  |  |  |  |  |  |
| Bisexual | **2.36** | **[1.93,2.89]** | **2.03** | **[1.66,2.49]** | **4.28** | **[3.31,5.53]** | **2.52** | **[1.94,3.27]** | **1.68** | **[1.21,2.32]** | 1.39 | [1.00,1.92] |
| Homosexual | **2.14** | **[1.83,2.50]** | **1.46** | **[1.23,1.73]** | **3.84** | **[2.93,5.03]** | **2.50** | **[1.91,3.28]** | **1.45** | **[1.21,1.75]** | 1.15 | [0.95,1.39] |
| **Heavy Drinking** |  |  |  |  |  |  |  |  |  |  |  |  |
| Bisexual | **2.52** | **[1.76,3.60]** | **2.06** | **[1.43,2.98]** | **4.15** | **[2.80,6.15]** | **2.53** | **[1.68,3.81]** | 1.38 | [0.61,3.10] | 1.16 | [0.51,2.63] |
| Homosexual | **2.35** | **[1.81,3.07]** | **1.68** | **[1.26,2.23]** | **4.17** | **[2.95,5.89]** | **3.36** | **[2.39,4.72]** | 1.14 | [0.82,1.58] | 0.91 | [0.65,1.29] |
| **Daily tobacco use** |  |  |  |  |  |  |  |  |  |  |  |  |
| Bisexual | 1.11 | [0.72,1.71] | 1.49 | [0.98,2.29] | 1.17 | [0.73,1.89] | **1.82** | **[1.12,2.96]** | 1.19 | [0.58,2.45] | 1.35 | [0.65,2.77] |
| Homosexual | **1.91** | **[1.51,2.41]** | **2.15** | **[1.71,2.69]** | **1.95** | **[1.27,2.99]** | **2.66** | **[1.74,4.07]** | **1.76** | **[1.32,2.34]** | **1.93** | **[1.45,2.56]** |
| **Multiple substance use/abuse** |  |  |  |  |  |  |  |  |  |  |  |  |
| Bisexual | **2.31** | **[1.28,4.17]** | **2.27** | **[1.27,4.07]** | **3.82** | **[2.06,7.09]** | **3.23** | **[1.72,6.05]** | 1.92 | [0.67,5.54] | 1.65 | [0.57,4.77] |
| Homosexual | **3.05** | **[2.14,4.34]** | **2.27** | **[1.60,3.23]** | **5.26** | **[2.71,10.19]** | **4.35** | **[2.35,8.07]** | **2.13** | **[1.39,3.27]** | **1.73** | **[1.14,2.62]** |

Note: heterosexual was the base category for the sexual orientation variable in the models. Models adjusted for: sex (only for total column), age groups, marital status, education level and household status. Prevalence ratios statistically significant are in boldface.
CPR, Crude Prevalence Ratio; CI, Confidence Interval; APR, Adjusted Prevalence Ratio.

Among women, regarding the adjusted estimates, only the specific prevalence ratio for depression among lesbians was not significant in comparison with heterosexuals. The estimates obtained for all outcomes, except for those for depression, were higher than those observed among men.

The estimated prevalence of heavy drinking among lesbians (APR=3.36, 95% CI: 2.39,4.72) and the use/abuse of multiple substances among bisexuals (APR=3.23, 95% CI: 1.72,6.05) were more than three times those observed among heterosexuals. While use/abuse of multiple substances among lesbians was more than four times (APR=4.35, 95% CI: 2.35, 8.07). The adjusted prevalence of depression, binge and heavy drinking among bisexuals, and binge drinking or daily tobacco use among lesbians were more than twice those observed among heterosexual women. Daily tobacco use among bisexuals was 1.82 (95% CI: 1.12, 2.96) times that observed among heterosexuals.

Among men, adjusted estimates for the prevalence of binge drinking and heavy drinking did not show significant differences between gays and bisexuals compared with heterosexuals. However, the prevalence of daily tobacco use (APR=1.93, 95% CI: 1.45;2.56) and use/abuse of multiple substances (APR=1.73, 95% CI 1.14;2.62) was significantly higher among gay men compared with heterosexuals.

With regard to mental health, we observed that the prevalence of depression among bisexuals (APR=2.91, 95% CI: 1.54,5.53) was almost three times higher than that observed among heterosexuals, while among gay men this ratio was close to twice (APR=1.92, CI95%: 1.32, 2.80).

## Discussion

Our study is the first to assess the association between sexual orientation and the presence of depression and substance use, according to sex, in one of the most important population-based health surveys in Brazil.

With respect to the differences between the profile of homosexuals and bisexuals compared with that of heterosexuals, our results are consistent with other findings, such as the fact that the LGB group is composed of younger and more educated individuals and has a higher proportion of single people and people residing in urban areas than among heterosexuals ^22,35^.

Regarding mental health, it is noteworthy that, compared with heterosexuals, bisexual individuals showed a prevalence of depression about two and a half times higher, with an even higher prevalence (three times) among bisexual men, than that observed among women. This finding may be related to the notion of monosexuality, biphobia or even the stigma attached to these individuals in the gay and lesbian community ^10,36,37^.

In a recent meta-analysis aimed at comparing the prevalence of depression and anxiety among bisexuals compared with heterosexuals and homosexuals ^38^, the authors found, among bisexuals compared with heterosexuals, a pooled odds ratio for the current depression outcome of 2.38 (95% CI: 1.86; 3.05), similar to our results, which also showed higher OR among men (OR=2.66, 95% CI 1.68; 4.21) than among women (OR=1.97, 95% CI: 1.40; 2.77).

Regarding substance use among women, a comparison between lesbians or bisexuals and heterosexuals showed significant differences for the prevalence ratios of all outcomes analyzed, with higher ratios than those observed among men.

Especially worrying is the prevalence of use/abuse of multiple substances, which is more than triple and almost four times higher among lesbians and bisexuals, respectively, compared with heterosexuals. According to Hughes et al. (2016), a possible explanation for this finding would be the non-acceptance by women belonging to sexual minorities of traditional gender expectations imposed by society, which are also reflected in substance use ^39^.

Among men, our study showed significant results related to the use/abuse of multiple substances in a comparison between homosexuals and heterosexuals. However, no significant differences were identified between gay or bisexual men and their heterosexual counterparts regarding alcohol abuse.

There are few studies that have assessed multiple substance use disparities related to sexual orientation. However, a populational-based study focused on adults from United States also found elevated and significantly odds ratio for multiple substance use disorders in the past year, among sexual minorities in 2004-2005. For bisexual and homosexual, the odds ratio was greater than four times those observed among heterosexual ^40^.

Furthermore, with regard to differences in excessive alcohol use among homosexuals and bisexuals compared with heterosexuals, according to sex, some population surveys focused on the US adult population corroborate our findings in this study.

In one of them, aimed at people aged 18 to 49, no difference was observed for binge drinking among gays of all age groups, as well as among bisexual men aged 26 to 49, while among lesbians aged 18 to 25 and bisexual women of all age groups analyzed the prevalence was higher compared with heterosexuals ^41^.

Another study, aimed at adults aged 18 and over, showed significant differences for binge drinking among lesbian and bisexual women compared with heterosexual women, while no such differences were identified among men ^42^.

### Strengths and limitations

This is the first time that sexual orientation has been investigated in one of the largest population-based health surveys in Brazil and methodological improvements are expected for future investigations. For this reason, our results should be interpreted with caution. Besides that, some estimates obtained, even if significant, showed low precision, with wide associated confidence intervals that should be taken in to account.

Sexual orientation was captured in the survey based on self-identification. Therefore, people who are attracted to or who engage in sexual activity with same-sex individuals, but who do not identify themselves as homosexuals or bisexuals, are included in the heterosexual group. This information is highly subject to underreporting and classification bias due to stigma and discrimination against the non-heteronormative population, especially in countries like Brazil, where conservatism and religious intolerance are still strong.

Despite the above issues, our findings are very consistent with those observed in other countries and, even if underestimated due to a possible classification bias, the effects found do point to the existence of important disparities between homosexuals and bisexuals relative to their heterosexual counterparts, both men and women.

Our study was limited to the investigation of sexual minorities in relation only to non-heteronormative sexual orientations and did not consider the part of the population that does not identify as cisgender, given that this information was not collected in the survey.

## Conclusion

Our study showed significant and worrying disparities regarding depression and harmful use of substances relative to sexual orientation among adults, pointing to relevant differences between the sexes that should be considered in future studies and in public policies aimed at minimizing both the negative social effects suffered by the non-heteronormative population and the existing disparities. We highlight the importance of paying greater attention to the health care needs of the sexual minority population. Interventions aimed at reducing tobacco use and alcohol abuse, especially among women, as well as the recognition and management of depression, especially among bisexuals, are extremely urgent.

## Data Availability

All data produced in the present work are contained in the manuscript

## Authors Contribution

N.L.G. conceptualized and designed the work, contributed to the acquisition, analysis, and interpretation of data for the work and drafted the article. C.S.L. edited and revised the article critically for important intellectual content.

All co-authors reviewed and approved the article before submission.

## References

1. Plöderl M, Tremblay P. Mental health of sexual minorities. A systematic review. International Review of Psychiatry 2015;27(5):367–385.

2. Gonzales G, Przedworski J, Henning-Smith C. Comparison of health and health risk factors between lesbian, gay, and bisexual adults and heterosexual adults in the United States: Results from the National Health Interview Survey. JAMA Intern Med 2016;176(9):1344–1351.

3. Rice CE, Vasilenko SA, Fish JN, et al. Sexual minority health disparities: an examination of age-related trends across adulthood in a national cross-sectional sample. Annals of Epidemiology 2019; 31:20–25.

4. Talley AE, Gilbert PA, Mitchell J, et al. Addressing gaps on risk and resilience factors for alcohol use outcomes in sexual and gender minority populations. Drug and Alcohol Review 2016;35(4):484–93.

5. Shokoohi M, Salway T, Ahn B, et al. Disparities in the prevalence of cigarette smoking among bisexual people: a systematic review, meta-analysis and meta-regression Tobacco Control 2021;30:e78–e86.

6. Li J, Berg CJ, Weber AA, et al. Tobacco Use at the Intersection of Sex and Sexual Identity in the U.S., 2007-2020: A Meta-Analysis. American Journal of Preventive Medicine 2020;60(3):415–424

7. Peralta RL, Victory E, Thompson CL. Alcohol use disorder in sexual minority adults: Age- and sex-specific prevalence estimates from a national survey, 2015-2017. Drug Alcohol Depend 2019; 1;205:107673.

8. Hequembourg AL, Blayney JA, Bostwick W, et al. Concurrent Daily Alcohol and Tobacco Use among Sexual Minority and Heterosexual Women. Subst Use Misuse 2020;55(1):66–78.

9. Roxburgh A, Lea T, de Wit J, Degenhardt L. Sexual identity and prevalence of alcohol and other drug use among Australians in the general population. Int J Drug Policy 2016;28:76–82.

10. Boyd CJ, Veliz PT, Stephenson R, et al. Severity of Alcohol, Tobacco, and Drug Use Disorders Among Sexual Minority Individuals and Their “Not Sure” Counterparts. LGBT Health 2019;6(1):15–22.

11. Meyer IH. Minority stress and mental health in gay men. J Health Soc Behav 1995;36(1):38–56.

12. Meyer IH. Prejudice, social stress, and mental health in lesbian, gay, and bisexual populations: conceptual issues and research evidence. Psychol Bull 2003 Sep;129(5):674–697.

13. Figueroa WS, Zoccola PM. Sources of Discrimination and Their Associations With Health in Sexual Minority Adults. J Homosex 2016 Jun;63(6):743–763.

14. Mays VM, Cochran SD. Mental health correlates of perceived discrimination among lesbian, gay, and bisexual adults in the United States. Am J Public Health 2001;91(11):1869–1876.

15. Lee JH, Gamarel KE, Bryant KJ, et al. Discrimination, Mental Health, and Substance Use Disorders Among Sexual Minority Populations. LGBT Health 2016;3(4):258–265.

16. Slater ME, Godette D, Huang B, et al. Sexual Orientation-Based Discrimination, Excessive Alcohol Use, and Substance Use Disorders Among Sexual Minority Adults. LGBT Health 2017;4(5):337–344.

17. McCabe SE, Bostwick WB, Hughes TL, et al. The relationship between discrimination and substance use disorders among lesbian, gay, and bisexual adults in the United States. Am J Public Health 2010;100(10):1946–1952.

18. Geary RS, Tanton C, Erens B, et al. Sexual identity, attraction and behaviour in Britain: The implications of using different dimensions of sexual orientation to estimate the size of sexual minority populations and inform public health interventions. PLoS One 2018;13(1):0189607.

19. OECD. Society at a Glance 2019: OECD Social Indicators. OECD Publishing: Paris; 2019. Available from: https://www.oecd.org/social/society-at-a-glance-19991290.htm [Last accessed: May/03/2021].

20. Shahab L, Brown J, Hagger-Johnson G, et al. Sexual orientation identity and tobacco and hazardous alcohol use: findings from a cross-sectional English population survey. BMJ Open 2017;7:015058.

21. Patrão AL, Almeida MC, Matos SMA, et al. Gender, sexual orientation and health behaviors in the ELSA-Brasil cohort. Cogent Social Sciences 2020, 6: 1787695.

22. Gonzales G, Henning-Smith C. Health Disparities by Sexual Orientation: Results and Implications from the Behavioral Risk Factor Surveillance System. J Community Health 2017; 42 (6), 1163–1172.

23. Diehl A, Pillon SC, Caetano R, et al. Violence and substance use in sexual minorities: Data from the Second Brazilian National Alcohol and Drugs Survey (II BNADS). Arch Psychiatr Nurs 2020;34(1):41–48.

24. Oliveira JMD, Mott L. Mortes violentas de LGBT+ no brasil – 2019: Relatório do Grupo Gay da Bahia. 1. ed.; Editora Grupo Gay da Bahia, Salvador 2020.

25. Ministério da Saúde. Política Nacional de Saúde Integral de Lésbicas, Gays, Bissexuais, Travestis e Transexuais; 2013. Available from: https://bvsms.saude.gov.br/bvs/publicacoes/politica_nacional_saude_lesbicas_gays.pdf [Last accessed April/29/2021].

26. Abade EAF, Chaves SCL, Silva GC de O. Saúde da população LGBT: uma análise dos agentes, dos objetos de interesse e das disputas de um espaço de produção científica emergente. Physis 2020; 300418–8.

27. Dunn TL, Gonzalez CA, Costa AB, et al. Does the minority stress model generalize to a non-U.S. sample? An examination of minority stress and resilience on depressive symptomatology among sexual minority men in two urban areas of Brazil. Psychology of Sexual Orientation and Gender Diversity 2014; 1(2), 117– 131.

28. Lawrenz P, Habigzang LF. Minority Stress, Parenting Styles, and Mental Health in Brazilian Homosexual Men. Journal of Homosexuality 2020; 67:5,658–673.

29. Stopa SR, Szwarcwald CL, Oliveira MM, et al. Pesquisa Nacional de Saúde 2019: histórico, métodos e perspectivas. Epidemiol. Serv. Saúde 2020; 29(5): 2020315;

30. IBGE. Pesquisa Nacional de Saúde 2019. Acidentes, violências, doenças transmissíveis, atividade sexual, características do trabalho e apoio social. Available from: https://www.ibge.gov.br/estatisticas/sociais/saude/9160-pesquisa-nacional-de-saude.html?edicao=30563&t=publicacoes [Last Accessed: May/15/2021].

31. Centers for Disease Control and Prevention (CDC). Excessive alcohol use. Available from: https://www.cdc.gov/chronicdisease/resources/publications/aag/alcohol.htm [Last Accessed: March/07/2021].

32. Moriarty AS, Gilbody S, McMillan D, Manea L. Screening and case finding for major depressive disorder using the Patient Health Questionnaire (PHQ-9): a meta-analysis. Gen Hosp Psychiatry 2015; 37:567–76.

33. Santos IS, Tavares BF, Munhoz TN A., Sensitivity and specificity of the Patient Health Questionnaire-9 (PHQ-9) among adults from the general population. Cad Saude Publica 2013;29(8),1533–43.

34. Barros A, Hirakata VN. Alternatives for logistic regression in cross-sectional studies: An empirical comparison of models that directly estimate the prevalence ratio. BMC Medical Research Methodology 2003; 3:21.

35. Pakula B, Shoveller J, Ratner PA, et al. Prevalence and Co-Occurrence of Heavy Drinking and Anxiety and Mood Disorders Among Gay, Lesbian, Bisexual, and Heterosexual Canadians. Am J Public Health 2016;106(6),1042–1048.

36. Bostwick W, Hequembourg AL. Minding the Noise: Conducting Health Research Among Bisexual Populations and Beyond. Journal of Homosexuality 2013; 60(4), 655–661.

37. Ross LE, Dobinson C, Eady A. Perceived determinants of mental health for bisexual people: a qualitative examination. Am J Public Health 2010;100(3):496–502.

38. Ross LE, Salway T, Tarasoff LA, et al. Prevalence of Depression and Anxiety Among Bisexual People Compared to Gay, Lesbian, and Heterosexual Individuals:A Systematic Review and Meta-Analysis. J Sex Res 2018;55(4-5),435–456.

39. Hughes TL, Wilsnack SC, Kantor LW. The Influence of Gender and Sexual Orientation on Alcohol Use and Alcohol-Related Problems: Toward a Global Perspective. Alcohol Res 2016;38(1):121–132.

40. McCabe SE, West BT. The 3-Year Course of Multiple Substance Use Disorders in the United States: A National Longitudinal Study. J Clin Psychiatry 2017;78(5):537–544.

41. Schuler MS, Rice CE, Evans-Polce RJ, et al. Disparities in substance use behaviors and disorders among adult sexual minorities by age, gender, and sexual identity. Drug Alcohol Depend 2018;189,139–146.

42. Fish JN, Hughes, TL, Russell, ST. Sexual identity differences in high-intensity binge drinking: findings from a US national sample. Addiction 2018; 113(4), 749– 758.

